# People with HIV receiving suppressive antiretroviral therapy show typical antibody durability after dual COVID-19 vaccination, and strong third dose responses

**DOI:** 10.1101/2022.03.22.22272793

**Authors:** Hope R. Lapointe, Francis Mwimanzi, Peter K. Cheung, Yurou Sang, Fatima Yaseen, Gisele Umviligihozo, Rebecca Kalikawe, Sarah Speckmaier, Nadia Moran-Garcia, Sneha Datwani, Maggie C. Duncan, Olga Agafitei, Siobhan Ennis, Landon Young, Hesham Ali, Bruce Ganase, F. Harrison Omondi, Winnie Dong, Junine Toy, Paul Sereda, Laura Burns, Cecilia T. Costiniuk, Curtis Cooper, Aslam H. Anis, Victor Leung, Daniel Holmes, Mari L. DeMarco, Janet Simons, Malcolm Hedgcock, Natalie Prystajecky, Christopher F. Lowe, Ralph Pantophlet, Marc G. Romney, Rolando Barrios, Silvia Guillemi, Chanson J. Brumme, Julio S.G. Montaner, Mark Hull, Marianne Harris, Masahiro Niikura, Mark A. Brockman, Zabrina L. Brumme

## Abstract

**Background:** Longer-term humoral responses to two-dose COVID-19 vaccines remain incompletely characterized in people living with HIV (PLWH), as do initial responses to a third dose.

**Methods:** We measured antibodies against the SARS-CoV-2 spike protein receptor-binding domain, ACE2 displacement and viral neutralization against wild-type and Omicron strains up to six months following two-dose vaccination, and one month following the third dose, in 99 PLWH receiving suppressive antiretroviral therapy, and 152 controls.

**Results:** Though humoral responses naturally decline following two-dose vaccination, we found no evidence of lower antibody concentrations nor faster rates of antibody decline in PLWH compared to controls after accounting for sociodemographic, health and vaccine-related factors. We also found no evidence of poorer viral neutralization in PLWH after two doses, nor evidence that a low nadir CD4+ T-cell count compromised responses. Post-third-dose humoral responses substantially exceeded post-second-dose levels, though anti-Omicron responses were consistently weaker than against wild-type.

Nevertheless, post-third-dose responses in PLWH were comparable to or higher than controls. An mRNA-1273 third dose was the strongest consistent correlate of higher post-third-dose responses.

**Conclusion:** PLWH receiving suppressive antiretroviral therapy mount strong antibody responses after two- and three-dose COVID-19 vaccination. Results underscore the immune benefits of third doses in light of Omicron.

## INTRODUCTION

As people living with HIV (PLWH) may be at increased risk of severe COVID-19 due to immunosuppression, higher rates of multi-morbidity and/or social determinants of health [1-4], COVID-19 vaccination is expected to benefit this group. Two-dose COVID-19 vaccination protects against severe disease [5-7], but impaired responses have been observed in certain immunocompromised groups [8-12]. While antiretroviral therapy can reverse HIV-induced immune dysfunction to a large extent [13-16], persistent HIV-related immunopathology can nevertheless blunt vaccine responses [17-19], prompting initial concern that PLWH may respond sub-optimally to COVID-19 immunization. Data from clinical trials [20, 21] and real-world studies however [22-27], including from our group [28], described strong initial immune responses to two-dose COVID-19 vaccination in PLWH with controlled HIV loads on therapy and preserved CD4+ T-cell counts [20-24, 28], though weaker responses have been observed in PLWH who are not receiving therapy or who have CD4+ T-cell counts <200 cells/mm^3^ [22, 25-27].

Vaccine-induced antibody responses decline over time, which can increase the risk of post-vaccination SARS-CoV-2 infection [29-31], particularly with the more transmissible Omicron variant [32-36]. Though immune response durability following two-dose COVID-19 vaccination has been examined among PLWH participants of the ChAdOx1 clinical trial [37], few real-world studies have investigated this. Furthermore, no studies to our knowledge have investigated immune responses in PLWH to third vaccine doses, despite their widespread recommendation to maintain protection [38-40]. Here, we extend our previous report [28] to characterize binding and neutralizing antibody responses up to six months following two-dose COVID-19 vaccination, as well as one month following the third dose, in 99 PLWH and 152 controls without HIV. We assess responses to both wild-type and Omicron SARS-CoV-2 variants.

## METHODS

### Participants

We previously recruited 99 adult PLWH and 152 controls without HIV, the latter predominantly health care workers, in British Columbia (BC), Canada [28]. Serum and plasma (collected in either ethylenediaminetetraacetic acid [EDTA] or anticoagulant citrate dextrose [ACD]) were collected before vaccination; one month after the first dose; one, three and six months after the second dose; and one month following the third dose. Specimens were processed same-day and frozen at −80°C until analysis. Here we report on the post-second- and third-dose time points.

### Ethics approval

All participants provided written informed consent. This study was approved by the University of British Columbia/Providence Health Care and Simon Fraser University Research Ethics Boards.

### Data sources

Sociodemographic, health and COVID-19 vaccine data were collected by self-report and confirmed through medical records where available. We assigned a score of 1 for each of 11 chronic conditions: hypertension; diabetes; asthma; obesity (body mass index ≥30 kg/m^2^); chronic diseases of lung, liver, kidney, heart or blood; cancer; and immunosuppression due to chronic conditions or medication. For PLWH, a recent CD4+ T-cell count <200 cells/mm^3^ constituted immunosuppression.

### Binding antibody assays

We measured total binding antibodies against SARS-CoV-2 nucleocapsid (N) and spike (S) receptor binding domain (RBD) in serum using the Elecsys Anti-SARS-CoV-2 and Anti-SARS-CoV-2 S assays, respectively, on a Cobas e601 module analyzer (Roche Diagnostics). Post-infection, both assays should be positive, whereas post-vaccination only the S assay should be positive. Both tests are electro-chemiluminescence sandwich immunoassays, and report results in arbitrary Units/mL. For the S assay, the manufacturer indicates that these arbitrary Unit (U) values can be considered equivalent to WHO-defined international binding antibody units [41]. For the S assay, sera were tested undiluted, with samples above the upper limit of quantification (ULOQ) re-tested at 1:100 dilution, allowing a measurement range of 0.4-25,000 U/mL. Anti-RBD binding IgG concentrations in serum were quantified using the V-plex SARS-CoV-2 (IgG) Panel 22 ELISA kit (Meso Scale Diagnostics), which features wild-type and Omicron antigens, on a Meso QuickPlex SQ120 instrument. Sera were diluted 1:10000, with results reported in arbitrary Units/mL.

### ACE2 displacement assay

We assessed the ability of serum antibodies to block the RBD-ACE2 receptor interaction by competition ELISA (Panel 22 V-plex SARS-CoV-2 [ACE2]; Meso Scale Diagnostics) on a Meso QuickPlex SQ120 instrument. Sera were diluted 1:40 and results reported as % ACE2 displacement.

### Live virus neutralization

Neutralizing activity in plasma was examined in live SARS-CoV-2 assays using isolate USA-WA1/2020 (BEI Resources) and a local Omicron BA.1 isolate (GISAID Accession # EPI_ISL_9805779) on VeroE6-TMPRSS2 (JCRB-1819) target cells. Viral stock was adjusted to 50 TCID_50_/200 μl in Dulbecco’s Modified Eagle Medium in the presence of serial 2-fold plasma dilutions (from 1/20 to 1/2560), incubated at 4°C for 1 hour and added to target cells in 96-well plates in triplicate. Cultures were maintained at 37°C with 5% CO_2_ and the appearance of viral cytopathic effect (CPE) was recorded three days post-infection. Neutralizing activity is reported as the reciprocal of the highest plasma dilution able to prevent CPE in all triplicate wells. Samples exhibiting partial or no neutralization at 1/20 dilution were assigned a reciprocal dilution of 10, defined as below the limit of quantification (BLOQ).

### Statistical analysis

Continuous variables were compared using the Mann-Whitney U-test (unpaired data) or Wilcoxon test (paired data). Relationships between continuous variables were assessed using Spearman’s correlation. Multiple linear regression was used to investigate the relationship between sociodemographic, health and vaccine variables and immune outcomes, except for neutralization at 6 months post-second dose, where multiple logistic regression was used due to the high proportion of results BLOQ. Variables included HIV infection (controls as reference group), age (per year), sex at birth (female as reference), ethnicity (non-white as reference), number of chronic conditions (per additional), interval between first and second doses (per day), sampling date after vaccination (per day), dual ChAdOx1 as the initial regimen (mRNA or mixed [ChAdOx1/mRNA] regimen as the combined reference group), and prior COVID-19 (COVID-19-naive as reference). Plasma neutralization models also corrected for the anticoagulant (ACD as reference). Post-third-dose analyses also corrected for the third dose mRNA vaccine brand (BNT162b2 as reference) and the interval between second and third doses (per day). All tests were two-tailed, with p<0.05 considered statistically significant. Analyses were conducted using Prism v9.2.0 (GraphPad).

## RESULTS

### Cohort characteristics

All PLWH were receiving antiretroviral therapy and had suppressed plasma HIV loads (**Table 1**). Recent CD4+ T-cell counts, measured a median 44 days before enrolment, were a median 715 cells/mm^3^ (Interquartile Range [IQR] 545-943; range 130-1800), where only two participants had values <200 cells/mm^3^. Nadir CD4+ T-cell counts were a median 280 cells/mm^3^ (IQR 123-490; range <10-1010). The 99 PLWH and 152 controls were broadly similar in age, but the PLWH group included a greater proportion of males and of white ethnicity. PLWH and controls had similar numbers of chronic health conditions (45% and 33%, respectively, had at least one condition). At study entry, 8% of PLWH and 10% of controls had anti-N antibodies, indicating prior SARS-CoV-2 infection. An additional 31 participants (18 PLWH; 13 controls) experienced post-vaccination SARS-CoV-2 infections, 26 of which occurred during the Omicron wave. More PLWH received dual ChAdOx1 vaccines for their first two doses (8%) compared to <1% of controls. On average, the interval between first and second doses was longer for controls (median 89 days versus 58 for PLWH). In British Columbia (BC), third doses began to be offered in October 2021 to priority populations, including PLWH who had one or more of: age ≥ 65 years, prior AIDS-defining illness, prior CD4 count <200 cells/mm^3^ or prior CD4 fraction ≤ 15%, any plasma HIV load >50 copies/mL in 2021, or perinatally-acquired HIV. The majority of PLWH in BC met at least one of these criteria. By January 2022, all remaining adults in BC aged ≥18 years were eligible for a third dose 6 months after their second dose. At the time of writing, 80% of PLWH participants and 88% of controls had received a third dose, on average 6.3 months following their second dose. All third doses were mRNA vaccines, and more PLWH (70%) received mRNA-1273 compared to controls (59%). Third mRNA-1273 doses also differed by risk group: 100 mcg was recommended for adults aged ≥70 years and PLWH who met any priority criterion, whereas the standard booster dose of 50 mcg was recommended for all other adults.

**Table 1:** Participant characteristics.

| Characteristic | PLWH (n=99) | Controls (n=152) |
| --- | --- | --- |
| **HIV-related variables** |  |  |
| Receiving antiretroviral therapy, n (%) | 99 (100%) | - |
| Most recent plasma viral load, copies HIV RNA/mL, median [IQR] | <50 [<50 - <50] | - |
| Most recent CD4+ T-cell count in cells/mm3, median [IQR] | 715 [545-943] | - |
| Nadir CD4+ T-cell count in cells/mm3, median [IQR] | 280 [123-490] | - |
| **Sociodemographic and health variables** |  |  |
| Age in years, median [IQR] | 54 [40-61] | 47 [35-70] |
| Male sex at birth, n (%) | 87 (88%) | 50 (33%) |
| Ethnicity, n (%) |  |  |
| white/Caucasian | 69 (69%) | 78 (51%) |
| Black | 5 (5%) | 1 (0.7%) |
| Asian | 10 (10%) | 59 (38%) |
| Latin American | 8 (8%) | 4 (2.6%) |
| Middle Eastern/Arab | 3 (3%) | 0 (0%) |
| Mixed ethnicity | 3 (3%) | 8 (5.3%) |
| Not disclosed | 1 (1%) | 2 (1.3%) |
| Number of chronic health conditions, median [IQR] | 0 [0-1] | 0 [0-1] |
| Hypertension, n (%) | 15 (15%) | 22 (14.5%) |
| Diabetes, n (%) | 6 (6%) | 6 (3.9%) |
| Asthma, n (%) | 7 (7%) | 15 (9.9%) |
| Obesity, n (%) | 15 (15%) | 14 (9.2%) |
| Chronic lung disease, n (%) | 4 (4%) | 3 (2%) |
| Chronic liver disease, n (%) | 4 (4%) | 1 (0.7%) |
| Chronic kidney disease, n (%) | 1 (1%) | 1 (0.7%) |
| Chronic heart disease, n (%) | 1 (1%) | 4 (2.6%) |
| Chronic blood disease, n (%) | 1 (1%) | 2 (1.3%) |
| Cancer, n (%) | 4 (4%) | 4 (2.6%) |
| Immunosuppression, n (%) | 3 (3%) | 0 (0%) |
| At least one of the above, n (%) | 45 (45%) | 50 (33%) |
| **COVID-19 status** |  |  |
| COVID-19 convalescent (anti-N Ab+) at study entry, n (%) | 8 (8%) | 15 (10%) |
| COVID-19 post-vaccination | 18 (18%) | 13 (9%) |
| **Vaccine details** |  |  |
| Initial two-dose regimen |  |  |
| mRNA - mRNA | 82 (82%) | 148 (97%) |
| ChAdOx1 - mRNA (heterologous) | 8 (8%) | 3 (2%) |
| ChAdOx1 - ChAdOx1 | 8 (8%) | 1 (0.7%) |
| ChAdOx1 - not disclosed | 1 (1%) | - |
| Time between first and second doses in days, median [IQR] | 58 [53-67] | 89 [65-98] |
| Third dose |  |  |
| BNT162b2 | 23 of 80 (29%) | 55 of 134 (41%) |
| mRNA-1273 | 56 of 80 (70%) | 79 of 134 (59%) |
| Unknown | 1 of 80 (1%) | - |
| Time between second and third doses in days, median [IQR] | 183 [143-191] | 198 [173-216] |
| **Specimen collection** |  |  |
| Specimen one month after second dose, n (%) | 97 (97%) | 151 (99%) |
| Day of collection one month after second dose, median [IQR] | 30 [29-30] | 30 [29-32] |
| Specimen three months after second dose, n (%) | 96 (96%) | 148 (97%) |
| Day of collection three months after second dose, median [IQR] | 90 [90-91] | 90 [89-91] |
| Specimen six months after second dose, n (%) | 62 (62%) | 136 (89%) |
| Day of collection six months after second dose, median [IQR] | 180 [177-182] | 180 [178-182] |
| Specimen one month after third dose, n (%) | 80 (80%) | 134 (88%) |
| Day of collection one month after third dose, median [IQR] | 30 [30-32] | 30 [29-32] |

### Binding antibody responses after second and third doses

One month following two-dose vaccination, anti-RBD antibody concentrations were a median 3.9 [IQR 3.7-4.1] log_10_ U/mL in PLWH compared to a median of 4.0 [IQR 3.8-4.2] log_10_ in controls (p=0.04, **Figure 1A**). By three months following the second dose, antibody concentrations had declined in both groups, to a median of 3.4 [IQR 3.2-3.6] log_10_ U/mL in PLWH compared to a median of 3.6 [IQR 3.4-3.8] log_10_ U/mL in controls (p=0.0001). These differences however did not remain significant in multivariable analyses controlling for sociodemographic, health- and vaccine-related variables (p-values for HIV infection p=0.83 and p=0.088, respectively, **Supplemental Table 1**). Rather, a greater number of chronic conditions and dual ChAdOx1 vaccination were independently associated with lower antibody concentrations at both time points, while a longer dose interval was associated with higher antibody concentrations (all p<0.05), regardless of HIV status. Older age was also significantly correlated with lower antibody concentrations one month post-second dose (p=0.0053), and was marginally significant at three months (p=0.055). Participants with prior COVID-19 (where post-vaccination infections are shown as red dots on **Figure 1A**) displayed modestly higher responses at one- and three-months post-second dose, though this did not remain significant after multivariable correction.

**Figure 1.**
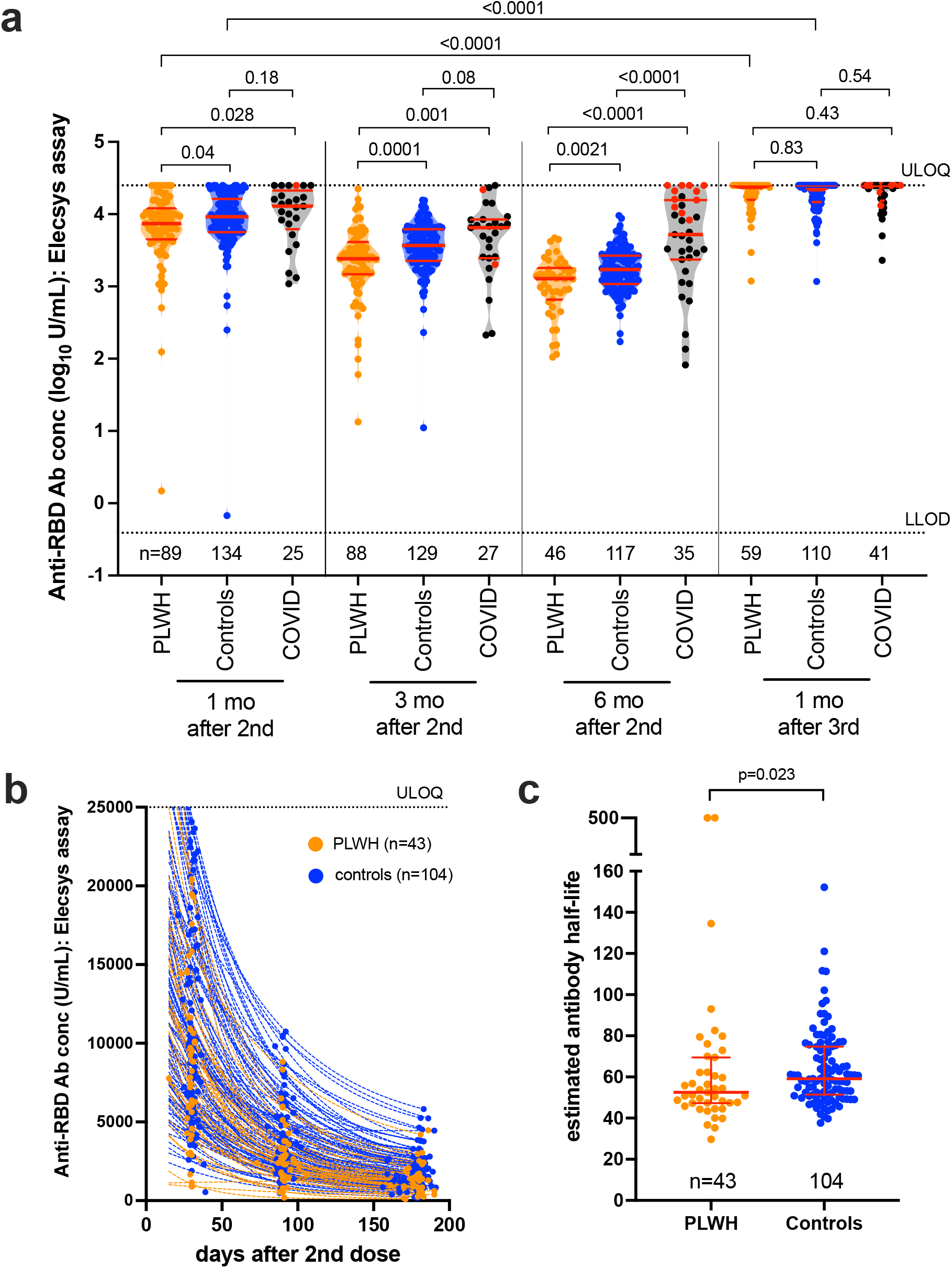
Concentrations of total binding antibodies in serum to spike RBD following two and three COVID-19 vaccine doses. *Panel A:* Binding antibody responses to the SARS-CoV-2 spike RBD in serum at one, three and six months following the second dose, and one month following the third vaccine dose, in PLWH (orange) and controls (blue) who were COVID-19 naive at the studied time point, as well as individuals who had recovered from COVID-19 at the studied time point (COVID group, black). Participants who experienced a post-vaccination infection were relocated from their original group into the COVID group at their first post-infection study visit, where they are denoted by a red symbol. Participant Ns are shown at the bottom of the plot. The thick horizontal red bar represents the median; thinner horizontal red bars represent the IQR. P-values were computed using the Mann-Whitney U-test (for comparisons between groups) or the Wilcoxon matched pairs test (for comparisons across time points within a group) and are uncorrected for multiple comparisons. ULOQ: upper limit of quantification; LLOD: lower limit of detection. *Panel B:* Temporal declines in serum binding antibody responses to spike-RBD following two vaccine doses in PLWH (orange) and controls (blue) who remained COVID-19 naive during this period. Only participants with a complete longitudinal data series with no values above the ULOQ are shown. *Panel C*: Binding antibody half-lives following two COVID-19 vaccine doses, calculated by fitting an exponential curve to the data shown in panel B. Ns are indicated at the bottom of the plot. Red bars and whiskers represent the median and IQR. P-value computed using the Mann-Whitney U-test.

By six months after the second dose, antibody concentrations had declined to a median of 3.1 [IQR 2.9-3.3] log_10_ U/mL in PLWH versus a median 3.2 [IQR 3.0-3.4] log_10_ U/mL in controls (p=0.0021, **Figure 1A**), though this difference did not remain significant after multivariable correction (p=0.64; **Table 2**). Rather, dual ChAdOx1 vaccination was the strongest correlate of weaker responses at this time point, being associated with a nearly a log_10_ adjusted lower antibody concentration (p<0.0001), regardless of HIV status. Age was no longer a correlate of weaker responses at the six-month time point (p=0.99), while a longer time between vaccination and sampling was associated with marginally higher antibody concentrations (p=0.0067). This is likely driven by 13 control participants aged ≥ 70 years who did not contribute samples to this time point due to receipt of third doses less than six months after the second dose, and 25 participants aged ≥ 65 years who contributed this sample early due to imminently scheduled third doses as per the age-based rollout in BC. Prior COVID-19 was associated with superior antibody concentrations at the six-month time point (**Figure 1A** and **Table 2**), though this is influenced by 11 participants with recent infections.

**Table 2:** Multivariable analyses of the relationship between sociodemographic, health and vaccine-related variables on antibody concentrations 6 months after the second dose, and antibody half-lives following the second dose.

| Variable<sup>a</sup> | Log<sub>10</sub> antibody concentration 6 mo after 2nd dose<sup>b</sup> |  |  | Antibody half-lives after the 2nd dose<sup>c</sup> |  |  |
| --- | --- | --- | --- | --- | --- | --- |
|  | Estimate | 95% CI | p-value | Estimate | 95% CI | p-value |
| HIV infection | -0.036 | -0.19 to 0.11 | 0.64 | 6.33 | -19.92 to 32.59 | 0.63 |
| Age (per year) | 0.000019 | -0.0043 to 0.0043 | 0.99 | 0.53 | -0.12 to 1.17 | 0.11 |
| Male sex | -0.059 | -0.19 to 0.072 | 0.37 | 9.36 | -12.83 to 31.54 | 0.41 |
| White ethnicity | -0.0078 | -0.13 to 0.11 | 0.90 | -7.03 | -26.97 to 12.90 | 0.49 |
| # Chronic conditions (per additional) | -0.028 | -0.11 to 0.056 | 0.51 | -7.58 | -21.67 to 6.51 | 0.29 |
| Dual ChAdOx1 as initial regimen | -0.94 | -1.39 to -0.49 | **<0.0001** | 2.84 | -76.02 to 81.70 | 0.94 |
| Interval between first and second doses (per day) | 0.0024 | -0.00036 to 0.0052 | 0.087 | -0.17 | -0.63 to 0.29 | 0.47 |
| Days since second dose | 0.012 | 0.0033 to 0.020 | **0.0067** | - | - | - |
| Prior COVID-19 | 0.50 | 0.35 to 0.65 | **<0.0001** | - | - | - |

<sup>a</sup> Dashes indicate variables not included in the model
<sup>b</sup> quantified using the Roche Elecsys anti-S assay
<sup>c</sup> Calculated from all participants with a complete longitudinal data series following the second dose with no values above the ULOQ, and no evidence of prior COVID-19

We next assessed temporal reductions in antibody concentrations after two vaccine doses (**Figure 1B)**. Assuming exponential decay, and restricting the analysis to COVID-19-naive participants with a complete post-second-dose longitudinal series with no values above the assay upper limit of quantification (ULOQ), we estimated antibody half-lives to be a median of 53 [IQR 47-70] days in PLWH versus a median of 59 [IQR 51-75] days in controls (p=0.023, **Figure 1C**). This difference however did not remain significant after multivariable correction (p=0.63; **Table 2**).

A third vaccine dose boosted antibody concentrations to an average of 0.4-0.5 log_10_ U/mL higher than peak post-second dose levels (within-group p<0.0001 for both PLWH and controls), to a median of 4.3 [IQR 4.2 to >ULOQ] log_10_ U/mL in PLWH and 4.4 [IQR 4.2 to >ULOQ] log_10_ U/mL in controls (between-group p=0.83), values that were comparable to those in participants with prior COVID-19 (**Figure 1A**). Multivariable analyses were not performed as nearly 50% of values were >ULOQ.

Consistent with our previous observations at one and three months post-second vaccine dose [28], we observed no significant relationship between most recent or nadir CD4+ T-cell count and antibody concentrations either six months after the second dose or one month following the third dose in PLWH (**Supplementary Figure 1**). We also observed no significant relationship between these CD4 parameters and antibody half-life after the second dose (Spearman ρ≤0.16, p≥0.3; not shown).

### Viral neutralization after second and third doses

One month after the second vaccine dose, SARS-CoV-2 neutralization was achieved at a median reciprocal plasma dilution of 160 (IQR 40-320) in PLWH compared to a median of 80 (IQR 40-160) in controls (Mann-Whitney p=0.06, **Figure 2A**). By three months post-second dose this activity declined to 40 [IQR 20-80] in both PLWH and controls (p=0.23). Multivariable analyses identified older age, a higher number of chronic conditions and dual ChAdOx1 vaccination as significant independent correlates of poorer neutralization one month post-second dose (all p<0.05), with negative effects of dual ChAdOx1 vaccination (p=0.0032) and to a lesser extent age (p=0.059) persisting at three months (**Supplemental Table 1**). Prior COVID-19 was associated with higher neutralization at both of these time points following multivariable correction (both p≤0.0002).

**Figure 2.**
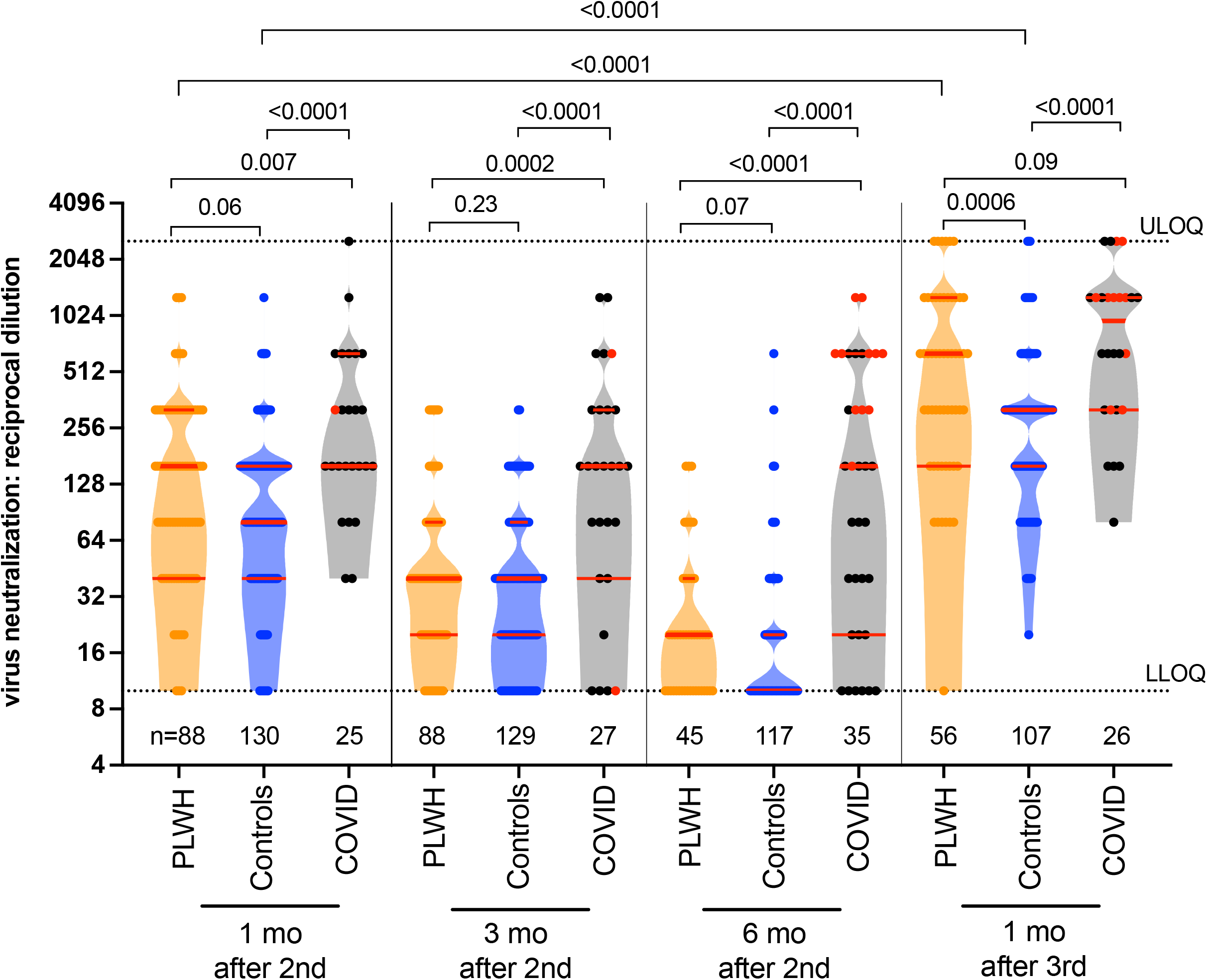
Live virus neutralization activities following two and three COVID-19 vaccine doses. Viral neutralization activity in plasma at one, three and six months following the second dose, and one month following the third vaccine dose, in PLWH (orange) and controls (blue) who were COVID-19 naive at the studied time point, as well as individuals who had recovered from COVID-19 at the studied time point (COVID group, black). Plasma neutralization was defined as the reciprocal of the highest plasma dilution at which vial cytopathic effect was prevented in all triplicate assay wells. Plasma samples showing neutralization in fewer than three wells at the lowest plasma dilution of 1/20 were coded as having a reciprocal dilution of 10, corresponding to the lower limit of quantification (LLOQ) in this assay. The highest dilution tested was 1/2560, which corresponds to the upper limit of quantification (ULOQ). Participants who experienced a post-vaccination infection were relocated from their original group into the COVID19 group at their first post-infection study visit, where they are denoted by a red symbol. Participant Ns are shown at the bottom of the plot. The thick horizontal red bar represents the median; thinner horizontal red bars represent the IQR. P-values were computed using the Mann-Whitney U-test (for comparisons between groups) or the Wilcoxon matched pairs test (for comparisons across time points within a group) and are uncorrected for multiple comparisons.

By six months post-second dose, neutralization had declined to below the limit of quantification (BLOQ) in 52% of COVID-19-naive participants, to a median reciprocal dilution of 20 [IQR BLOQ-40] in PLWH and a median BLOQ [IQR BLOQ-20] in controls (p=0.07, **Figure 2A**). Due to the large proportion of BLOQ values, we applied multivariable logistic regression with neutralization as a binary variable, and identified only prior COVID-19 as a biological correlate of neutralization at this time point (p=0.0037; **Supplemental Table 2**). This however is influenced by 11 participants with recent infections.

A third COVID-19 vaccine dose boosted neutralization to an average of fourfold higher than peak post-second-dose levels (within-group p<0.0001 for PLWH and controls; **Figure 2A**). In fact, neutralization activities in PLWH (median reciprocal dilution of 640 [IQR 160-1280]) exceeded those of controls (median of 320 [160-320]; Mann-Whitney p=0.0006) at this time point, though this did not remain significant following multivariable adjustment (p=0.15; **Supplemental Table 3**). Rather, having received mRNA-1273 as a third dose was the strongest independent correlate of better neutralization (p=0.0009). Prior COVID-19 was associated with better neutralization, though it is difficult to disentangle infection-from vaccine-induced responses due to a number of recent infections (red circles in **Figure 2A**).

We observed no significant relationship between most recent CD4+ T-cell count and neutralization at either six months post-second dose nor at one month post-third dose in COVID-19 naive PLWH; nor any relationship between nadir CD4+ T-cell count and neutralization at six months post-second dose (**Supplemental Figure 1**). An inverse relationship between nadir CD4+ T-cell count and neutralization one month after the third dose however was found (Spearman ρ= −0.28; p=0.04).

### Humoral responses against Omicron following two and three vaccine doses

To estimate the extent to which a third dose boosts protection against the now-dominant Omicron variant, we compared peak responses against wild-type and Omicron variants one month following the second and third doses. To avoid confounding by infection-induced immunity, we restricted this analysis to COVID-19-naive individuals. For both PLWH and controls, serum IgG concentrations capable of binding Omicron RBD were on average ∼0.6 log_10_ U/mL lower than those capable of binding wild-type RBD at both time points (all within-group comparisons p<0.0001; **Figure 3A**). Nevertheless, the third dose significantly boosted anti-Omicron IgG concentrations to an average of 0.3-0.5 log_10_ U/mL higher than those observed after two doses in both groups (within-group comparisons p<0.0001). One month post-second dose, anti-Omicron IgG concentrations were a median 4.12 [IQR 3.93-4.35] log_10_ U/mL in PLWH and a median of 4.28 [IQR 3.97-4.56] log_10_ U/mL in controls (p=0.04), but after three doses, these responses reached equivalence, with medians of 4.51 [IQR 4.26-4.93] log_10_ U/mL in PLWH versus 4.56 [IQR 4.24-4.74] log_10_ U/mL in controls (p=0.63). In fact, a multivariable analysis of Omicron-specific IgG concentrations after three doses identified HIV infection as being associated with an adjusted 0.36 log_10_ U/mL *higher* anti-Omicron IgG concentrations (p=0.0017; **Table 3**). Having received mRNA-1273 for the third dose, as well as longer interval between second and third doses, were also associated with higher anti-Omicron IgG responses (both p<0.05); male sex was associated with lower responses (p=0.032).

**Figure 3:**
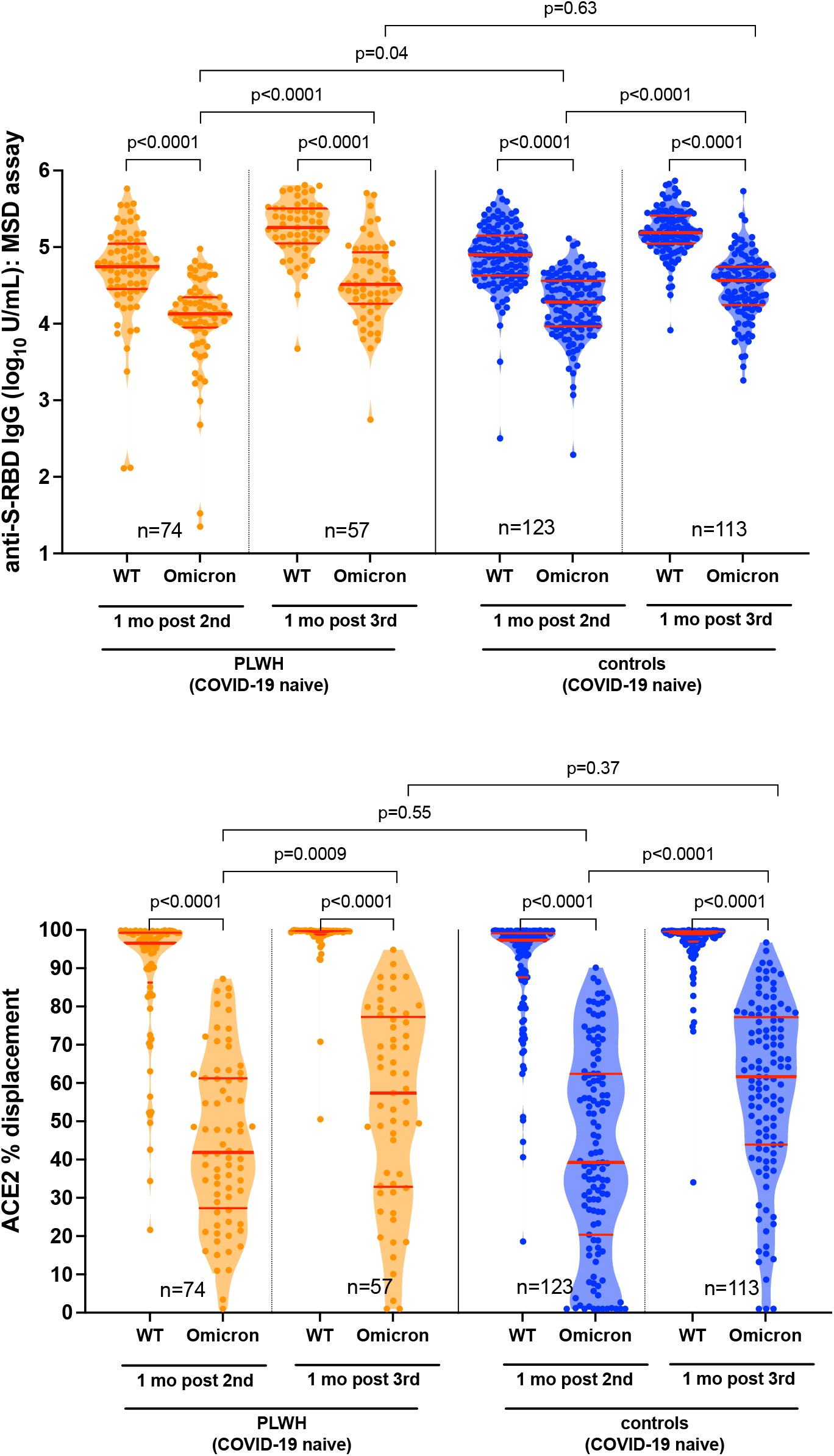
Anti-Omicron IgG binding and ACE2 displacement activities one month after the second and third COVID-19 vaccine doses. *Panel A:* Binding IgG responses in plasma to the wild-type (WT) and Omicron (OM) spike-RBD (S-RBD), measured using the Meso Scale Diagnostics V-Plex assay, in PLWH (orange) and controls (blue) who remained COVID-19 naive throughout the study. Participant Ns are shown at the bottom of the plot. Thick horizontal red bar represents the median; thinner horizontal red bars represent the IQR. P-values were computed using the Wilcoxon matched pairs test (for all within-group comparisons) or the Mann-Whitney U-test (for between-group comparisons) and are uncorrected for multiple comparisons. *Panel B:* same as A, but for ACE2 displacement activity, measured using the V-plex SARS-CoV-2 (ACE2) assay, where results are reported in terms of % ACE2 displacement.

**Table 3:** Multivariable analyses of the relationship between sociodemographic, health and vaccine-related variables on antibody responses to Omicron after three COVID-19 vaccine doses.

| Variable<sup>a</sup> | anti-Omicron log<sub>10</sub> Binding IgG<sup>b</sup> |  |  | anti-Omicron ACE2 % displacement<sup>b</sup> |  |  |
| --- | --- | --- | --- | --- | --- | --- |
|  | Estimate | 95% CI | p-value | Estimate | 95% CI | p-value |
| HIV infection | 0.36 | 0.14 to 0.58 | **0.0017** | 3.49 | -8.48 to 15.47 | 0.57 |
| Age (per year) | -0.0030 | -0.0078 to 0.0018 | 0.22 | 0.21 | -0.056 to 0.47 | 0.12 |
| Male sex | -0.19 | -0.36 to -0.017 | **0.032** | -10.26 | -19.58 to -0.94 | **0.031** |
| White ethnicity | 0.045 | -0.10 to 0.19 | 0.55 | 2.05 | -6.00 to 10.09 | 0.62 |
| # Chronic conditions (per additional) | 0.0032 | -0.083 to 0.090 | 0.94 | -1.27 | -5.99 to 3.45 | 0.60 |
| Dual ChAdOx1 as initial regimen (vs mixed or mRNA) | -0.14 | -0.48 to 0.20 | 0.42 | -6.80 | -25.39 to 11.80 | 0.47 |
| mRNA-1273 as third dose (vs BNT162b2) | 0.15 | 0.0074 to 0.30 | **0.039** | 5.16 | -2.80 to 13.12 | 0.20 |
| Interval between 1st and 2nd dose (per day) | 0.0022 | -0.0016 to 0.0059 | 0.26 | -0.044 | -0.25 to 0.16 | 0.67 |
| Interval between 2nd and 3rd dose (per day) | 0.0036 | 0.0012 to 0.0060 | **0.0039** | 0.065 | -0.067 to 0.20 | 0.33 |
| Days since 3rd dose | -0.0073 | -0.022 to 0.0080 | 0.35 | 0.11 | -0.72 to 0.94 | 0.80 |

<sup>a</sup> Analysis was restricted to participants with no evidence of prior COVID-19
<sup>b</sup> Both immunogenicity measures were quantified in serum using the Meso Scale Diagnostics V-Plex assay (panel 22) which features wild-type and Omicron S-RBD.

We also assessed the ability of plasma to block the RBD-ACE2 interaction, which estimates potential viral neutralization [42]. This activity was significantly weaker against Omicron compared to wild-type for both groups at both time points (all within-group comparisons p<0.0001; **Figure 3B**), where the discrepancy was most pronounced after two doses (*e*.*g*. median activities against wild-type and Omicron in PLWH were 97% versus 42%, respectively, at this time). The third dose nevertheless universally boosted anti-Omicron responses to above second-dose levels (all within-group comparisons p≤0.0009), with median anti-Omicron activity in PLWH rising from 42% after two doses to 57% after three (p=0.0009). Anti-Omicron ACE2 % displacement activities were comparable between groups at both time points: one month after the second dose these were a median 42% [IQR 27-61] in PLWH compared to 39% [IQR 20-62] in controls (p=0.55), rising to a median 57% [IQR 33-77] in PLWH compared to 62% [IQR 44-77] in controls one month after the third dose (p=0.37). In multivariable analyses, male sex was the only independent (negative) correlate of anti-Omicron ACE2 displacement activity after three doses (p=0.031, **Table 3**). After three doses, we observed a weak inverse relationship between nadir CD4+ T-cell count and anti-Omicron ACE2 % displacement (Spearman ρ= −0.3; p=0.02), but no relationship between other CD4+ T-cell count measures and anti-Omicron responses (**Supplemental Figure 1)**.

Finally, we assessed plasma neutralization against live wild-type and Omicron viruses at one month following the second and third doses in a subset of COVID-19-naive participants (**Figure 4**). While neutralization against Omicron was significantly weaker compared to wild-type at both time points in both PLWH and controls (all within-group comparisons p<0.0001), the third dose nevertheless significantly boosted anti-Omicron neutralization above second dose levels (both within-group comparisons p<0.0001). One month after the second dose, both PLWH and controls neutralized Omicron at a median reciprocal dilution of 20 [IQR BLOQ - 40] (p=0.71). One month after the third dose, anti-Omicron neutralization activity increased to a median reciprocal dilution of 80 [IQR 40-160] in PLWH compared to a median 40 [IQR 40-80] in controls (p=0.03). This was consistent with the superior neutralization of wild-type virus observed in PLWH at this timepoint (**Figure 2**). Neutralization of wild-type and Omicron viruses correlated significantly with their respective ACE2 displacement activities (all p<0.0001, **Supplemental Figure 2**).

**Figure 4:**
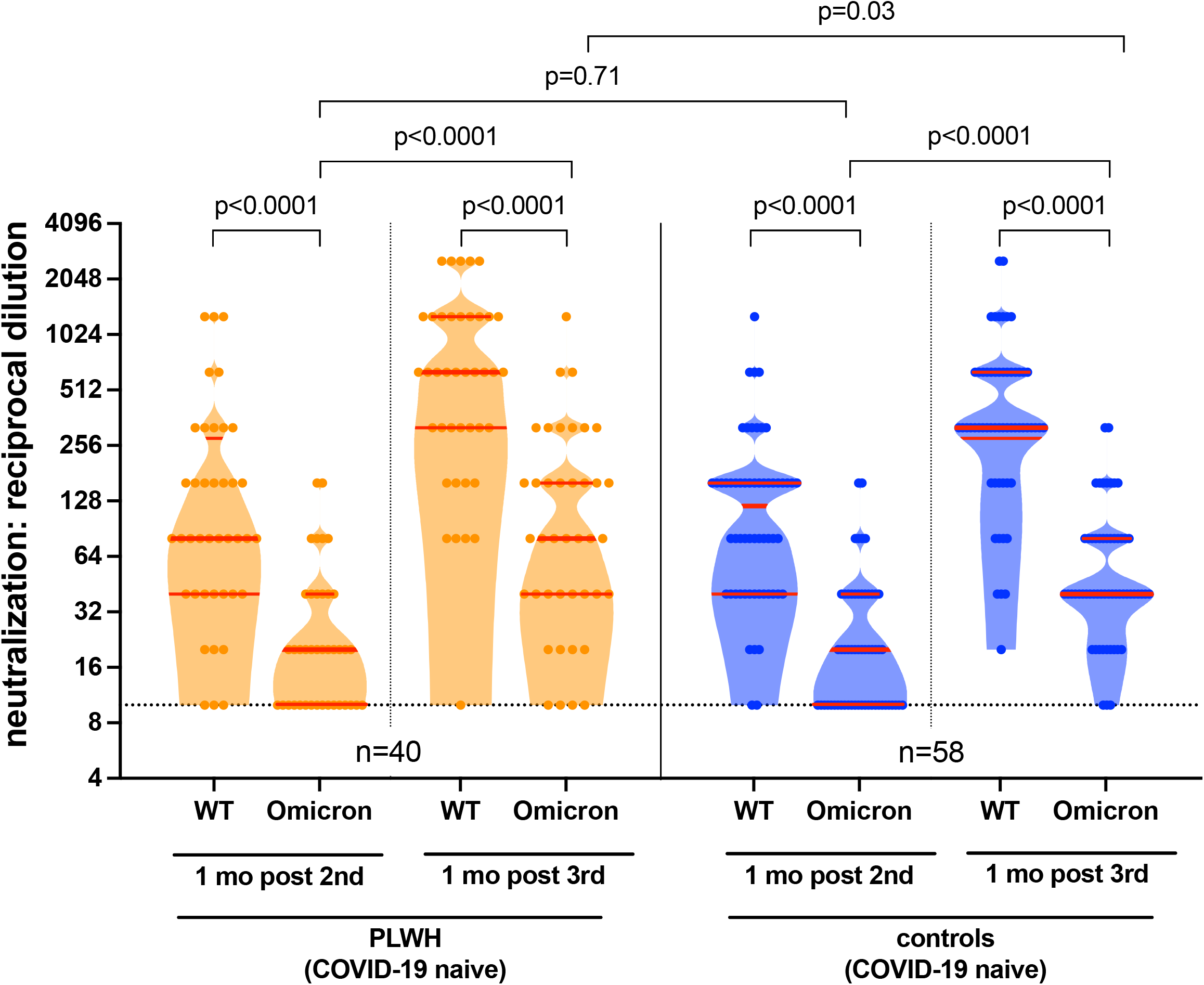
Anti-Omicron neutralization activities one month after the second and third COVID-19 vaccine doses. Neutralization activities, reported as the reciprocal of the highest plasma dilution at which neutralization was observed in all triplicate assay wells, against the wild-type (WT) and Omicron (OM) virus isolates a subset of PLWH (orange) and controls (blue) who remained COVID-19 naive throughout the study. Participant Ns are shown at the bottom of the plot. Thick horizontal red bar represents the median; thinner horizontal red bars represent the IQR. P-values were computed using the Wilcoxon matched pairs test (for within-group comparisons) or the Mann-Whitney U-test (for between-group comparisons) and are uncorrected for multiple comparisons.

## DISCUSSION

Our study confirms that antibody concentrations and neutralizing activities naturally decline following two-dose COVID-19 vaccination [31, 43]. Nevertheless, we found no evidence that PLWH receiving suppressive antiretroviral therapy exhibited lower antibody concentrations at any time point up to six months following two-dose vaccination, nor did they exhibit faster rates of antibody decline during this period compared to controls, after accounting for sociodemographic, health- and vaccine-related factors. Similarly we found no evidence that PLWH exhibited poorer neutralization at any time point after two doses compared to controls. These observations are consistent with data from PLWH participants of the original ChAdOx1 trial, which reported no significant difference in immune response decline in PLWH compared to controls following two vaccine doses [37].

Our results also showed that a third vaccine dose boosted binding antibody concentrations and function to significantly higher levels than those observed after two doses. After three doses, antibody concentrations in PLWH were equivalent to controls, while neutralization activities were slightly higher. The higher neutralization is attributable to PLWH more frequently receiving mRNA-1273 (vs. BNT162b2) third doses, which was the strongest correlate of higher neutralization after three-dose vaccination (**Supplemental Table 3**). In fact, the majority of PLWH were eligible for full (100 mcg) mRNA-1273 third doses, which likely boosted responses still further, though we were not able to confirm this due to incomplete dose information. Consistent with accumulating evidence [44-47], antibody responses against Omicron were universally weaker than against wild-type after two and three vaccine doses, though the third dose significantly boosted anti-Omicron responses. Indeed, post-third-dose anti-Omicron responses in PLWH were equivalent to or higher than controls, again possibly attributable to a higher proportion of PLWH receiving (full) mRNA-1273 third doses.

Our study has several limitations. Our results may not be generalizable to PLWH who are not receiving antiretroviral therapy, who have multiple co-morbidities or who have CD4+ T-cell counts <200 cells/mm^3^. We found no evidence that a low nadir CD4+ T-cell count negatively influenced COVID-19 vaccine response however, indicating that prior low CD4 T+ cell counts will not necessarily compromise immune responses to COVID-19 vaccines presently. We did not investigate T-cell responses, which may play critical protective roles, particularly against variants [48, 49]. Individuals ≥70 years old and PLWH meeting priority criteria were eligible for full mRNA-1273 third doses, but we could not directly assess mRNA-1273 dose-related effects on immune responses due to incomplete dosing information. Finally, while immune correlates of vaccine-mediated protection are being elucidated for SARS-CoV-2 [50], the implications of our results on individual-level protection from infection and disease remain uncertain, particularly in the context of Omicron.

In conclusion, adult PLWH with well-controlled viral loads and preserved CD4+ T-cell counts mount strong and functional antibody responses to two and three COVID-19 vaccine doses, including to Omicron, though it will be important to monitor these responses over time. Studies of PLWH who are not receiving antiretroviral treatment or who have low CD4+ T-cell counts are also needed.

## Data Availability

All data produced in the present study are available upon reasonable request to the authors

## ACKNOWLEDGEMENTS

We thank the leadership and staff of Providence Health Care for their support of this study. We thank the phlebotomists and laboratory staff at St. Paul’s Hospital, the BC Centre for Excellence in HIV/AIDS and Simon Fraser University for assistance. Above all, we thank the participants, without whom this study would not have been possible.

This work was supported by funding from Genome BC, the Michael Smith Foundation for Health Research, and the BCCDC Foundation for Public Health through a rapid SARS-CoV-2 vaccine research initiative in BC award (VAC-009 to ZLB, MAB). It was also supported by the Public Health Agency of Canada (PHAC) through two COVID-19 Immunology Task Force (CITF) COVID-19 Awards (to ZLB, MGR, MAB and to CTC, CC, AHA), the Canada Foundation for Innovation through Exceptional Opportunities Fund – COVID-19 awards (to CJB, CFL, MAB, MN, MLD, RP, ZLB), a British Columbia Ministry of Health–Providence Health Care Research Institute COVID-19 Research Priorities Grant (to CJB and CFL), the CIHR Canadian HIV Trials Network (CTN) (to AHA) and the National Institute of Allergy and Infectious Diseases of the National Institutes of Health (R01AI134229 to RP). MLD and ZLB hold Scholar Awards from the Michael Smith Foundation for Health Research. FA was supported by an SFU Undergraduate Research Award. GU and FHO are supported by Ph.D. fellowships from the Sub-Saharan African Network for TB/HIV Research Excellence (SANTHE), a DELTAS Africa Initiative [grant # DEL-15-006]. The DELTAS Africa Initiative is an independent funding scheme of the African Academy of Sciences (AAS)’s Alliance for Accelerating Excellence in Science in Africa (AESA) and supported by the New Partnership for Africa’s Development Planning and Coordinating Agency (NEPAD Agency) with funding from the Wellcome Trust [grant # 107752/Z/15/Z] and the UK government. The views expressed in this publication are those of the authors and not necessarily those of PHAC, CITF, AAS, NEPAD Agency, Wellcome Trust, the Canadian or UK governments or other funders.

## SUPPLEMENTAL FIGURES

**Supplemental Figure 1:**
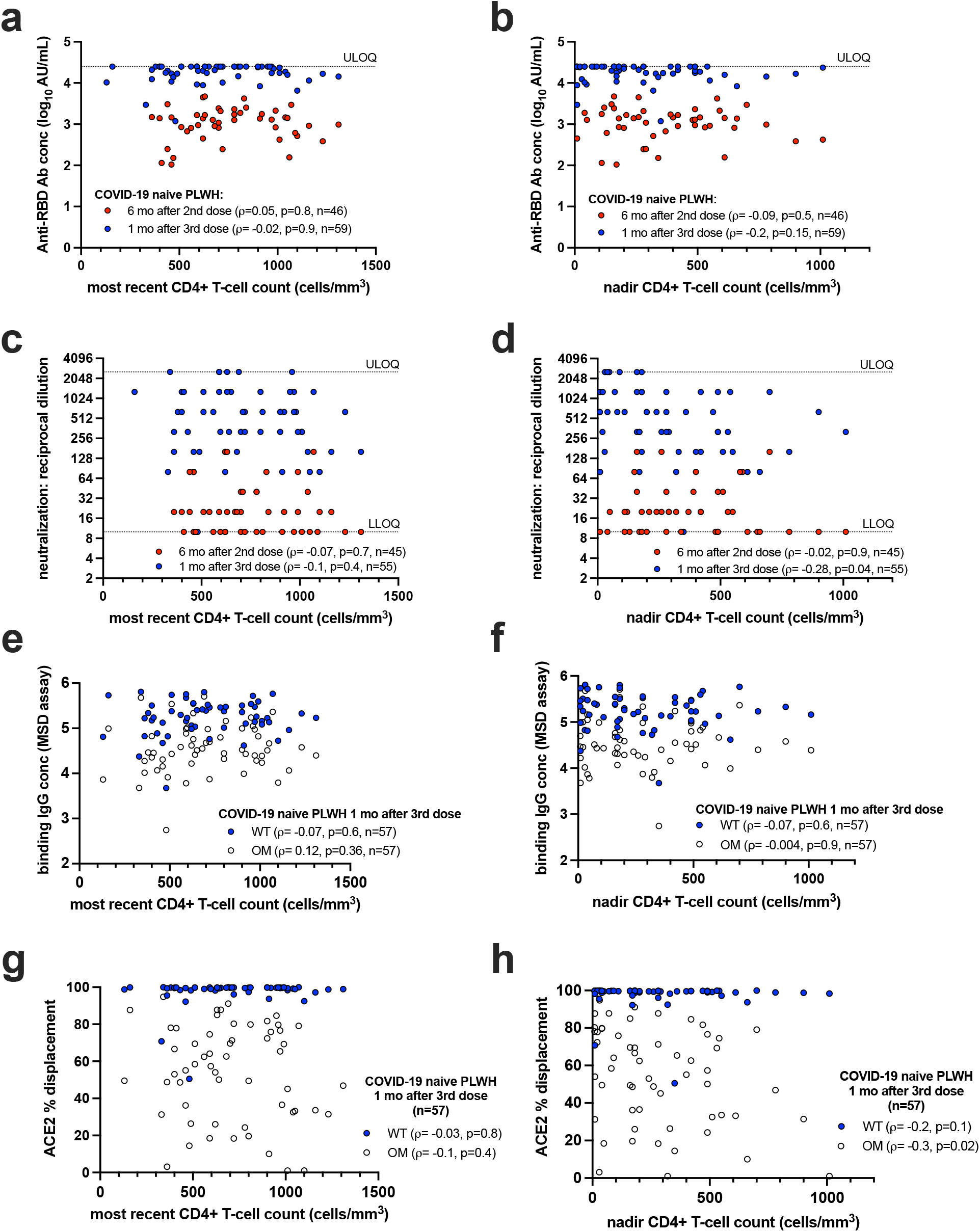
Relationships between most recent and nadir CD4+ T-cell counts and humoral responses following two and three vaccine doses. Relationships were assessed using Spearman’s correlation. Measurements against wild-type SARS-CoV-2 at six months post-second-dose are indicated by red symbols; measurements against wild-type SARS-CoV-2 at one month post-third-dose are indicated by blue symbols; measurements against Omicron at one month post-third-dose are indicated by open symbols. Analyses are restricted to COVID-19-naive PLWH. LLOQ: Lower limit of quantification; ULOQ: Upper limit of quantification.

**Supplemental Figure 2:**
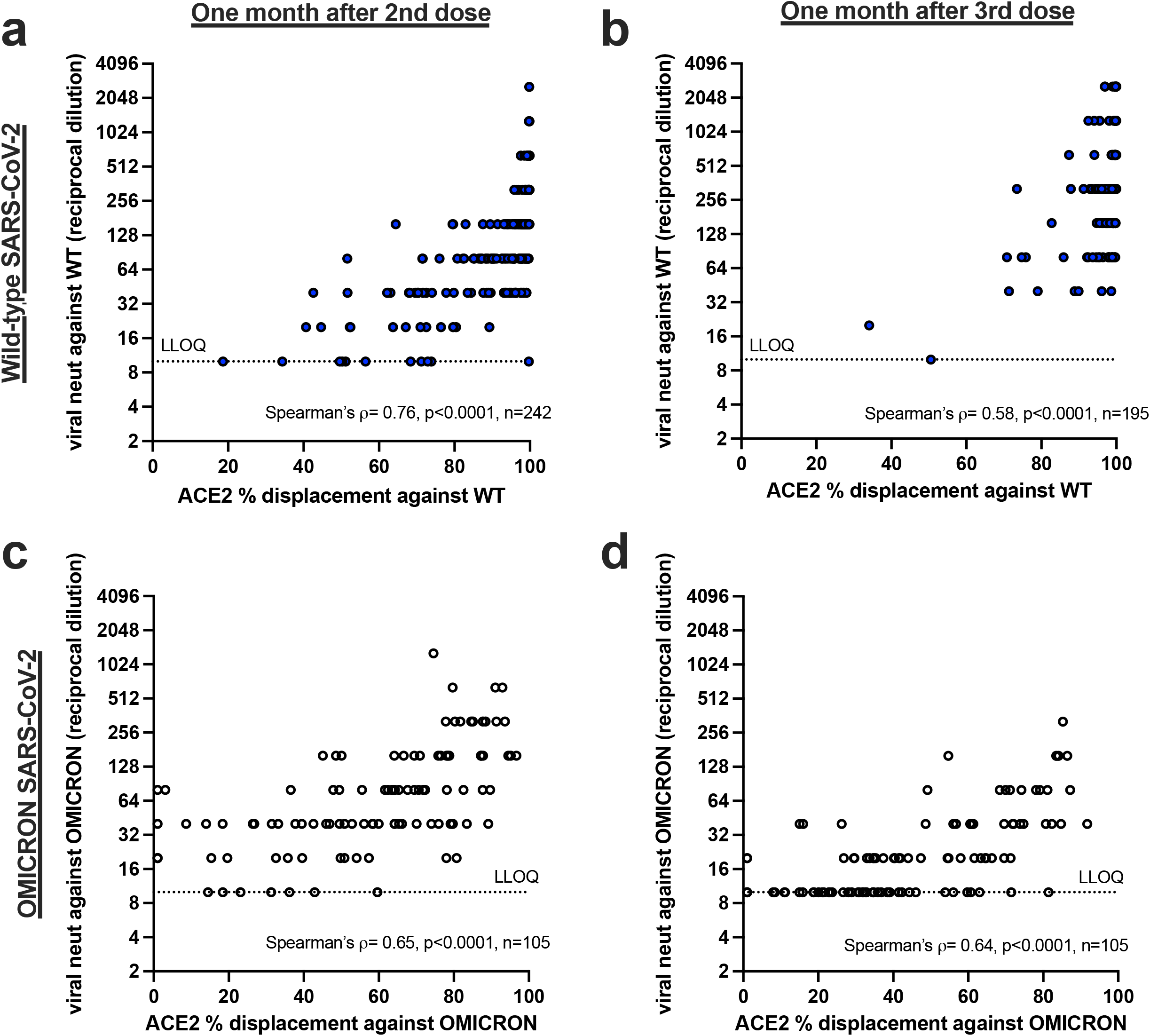
Relationships between ACE2 % displacement and viral neutralization activity against wild-type and Omicron after two and three COVID-19 vaccine doses. Relationships were assessed using Spearman’s correlation. Measurements against wild-type SARS-CoV-2 are indicated by blue symbols; measurements against Omicron are indicated by open symbols. Reported Ns reflect all measurements completed on all study participants (PLWH and controls) regardless of prior COVID-19 (while Figures 3 and 4 report only results from COVID-19-naive participants). LLOQ: Lower limit of quantification.

---

**Supplemental Table 1:** Multivariable analyses of the relationship between sociodemographic, health and vaccine-related variables on immunogenicity measures one and three months following the second vaccine dose.

| Immunogenicity outcome | Variable | Time point |  |  |  |  |  |
| --- | --- | --- | --- | --- | --- | --- | --- |
|  |  | 1 month after 2nd dose |  |  | 3 months after 2nd dose |  |  |
|  |  | Estimate | 95% CI | p-value | Estimate | 95% CI | p-value |
| **Log<sub>10</sub> anti-RBD Abs<sup>a</sup>** | HIV infection | -0.017 | -0.18 to 0.14 | 0.83 | -0.13 | -0.27 to 0.019 | 0.088 |
|  | Age (per year) | -0.0057 | -0.0098 to -0.0017 | **0.0053** | -0.0035 | -0.0072 to 0.000079 | 0.055 |
|  | Male sex | -0.016 | -0.16 to 0.13 | 0.82 | 0.028 | -0.10 to 0.16 | 0.68 |
|  | White ethnicity | 0.053 | -0.078 to 0.18 | 0.43 | 0.048 | -0.069 to 0.17 | 0.42 |
|  | # Chronic conditions (per additional) | -0.11 | -0.19 to -0.030 | **0.0072** | -0.086 | -0.16 to -0.012 | **0.022** |
|  | Dual ChAdOx1 as initial regimen | -0.63 | -0.97 to -0.29 | **0.0003** | -0.70 | -1.00 to -0.40 | **<0.0001** |
|  | Interval btw 1st and 2nd doses (per day) | 0.0036 | 0.00039 to 0.0069 | **0.028** | 0.0037 | 0.00085 to 0.0066 | **0.011** |
|  | Days since second dose | -0.0018 | -0.024 to 0.020 | 0.87 | 0.0061 | -0.010 to 0.022 | 0.46 |
|  | Prior COVID-19 | 0.063 | -0.13 to 0.26 | 0.53 | 0.14 | -0.030 to 0.31 | 0.10 |
| **Viral neutralization<sup>b</sup>** | HIV infection | 0.20 | -0.47 to 0.86 | 0.56 | -0.063 | -0.74 to 0.62 | 0.86 |
|  | Age (per year) | -0.018 | -0.030 to -0.0050 | **0.0064** | -0.012 | -0.025 to 0.00051 | 0.059 |
|  | Male sex | -0.39 | -0.84 to 0.055 | 0.086 | -0.12 | -0.59 to 0.34 | 0.60 |
|  | White ethnicity | -0.21 | -0.61 to 0.18 | 0.29 | -0.19 | -0.60 to 0.21 | 0.36 |
|  | # Chronic conditions (per additional) | -0.29 | -0.53 to -0.041 | **0.022** | -0.15 | -0.41 to 0.10 | 0.23 |
|  | Dual ChAdOx1 as initial regimen | -1.39 | -2.41 to -0.37 | **0.0077** | -1.57 | -2.60 to -0.53 | **0.0032** |
|  | Interval btw 1st and 2nd doses (per day) | 0.0073 | -0.0038 to 0.018 | 0.2 | 0.00067 | -0.010 to 0.012 | 0.91 |
|  | Days since second dose | -0.0061 | -0.073 to 0.060 | 0.86 | -0.029 | -0.084 to 0.027 | 0.31 |
|  | EDTA as anticoagulant<sup>c</sup> | 0.85 | 0.091 to 1.61 | **0.028** | 0.58 | -0.20 to 1.35 | 0.14 |
|  | Prior COVID-19 | 1.15 | 0.56 to 1.75 | **0.0002** | 1.65 | 1.06 to 2.24 | **<0.0001** |

<sup>a</sup>quantified using the Roche Elecsys anti-S assay
<sup>b</sup>for viral neutralization, reciprocal plasma dilutions were log<sub>2</sub> transformed prior to multivariable analysis.
<sup>c</sup>Neutralization assays were performed using plasma, so the analysis also corrects for the anticoagulant used, with ACD as the reference category. Analyses of anti-RBD concentration do not correct for this variable because this assay was performed on serum collected in the same tube type.

**Supplemental Table 2:** Multivariable analyses of the relationship between sociodemographic, health and vaccine-related variables and detectable viral neutralization activity six months following the second vaccine dose.

| Immunogenicity outcome | Variable | Odds Ratio | 95% CI | p-value |
| --- | --- | --- | --- | --- |
| Detectable viral neut. 6 months after 2nd dose<sup>a</sup> | HIV infection | 0.51 | 0.12 to 1.77 | 0.32 |
|  | Age (per year) | 0.98 | 0.96 to 1.01 | 0.22 |
|  | Male sex | 0.68 | 0.29 to 1.50 | 0.35 |
|  | White ethnicity | 0.88 | 0.44 to 1.78 | 0.73 |
|  | # Chronic conditions (per additional) | 1.00 | 0.61 to 1.63 | 1.00 |
|  | Dual ChAdOx1 as initial regimen | 0.19 | 0.0079 to 2.36 | 0.21 |
|  | Interval btw 1st and 2nd doses (per day) | 1.01 | 0.99 to 1.03 | 0.14 |
|  | Days since second dose | 1.03 | 0.98 to 1.09 | 0.24 |
|  | EDTA as anticoagulant<sup>b</sup> | 9.31 | 2.41 to 44.62 | **0.0023** |
|  | Prior COVID-19 | 4.57 | 1.74 to 13.99 | **0.0037** |

<sup>a</sup> Results are presented as the adjusted Odds Ratios and 95% CI of detectable viral neutralization activity at this time point, calculated using multivariable logistic regression.
<sup>b</sup> Neutralization assays were performed using plasma, so the analysis also corrects for the anticoagulant used, with ACD as the reference category.

**Supplemental Table 3:**
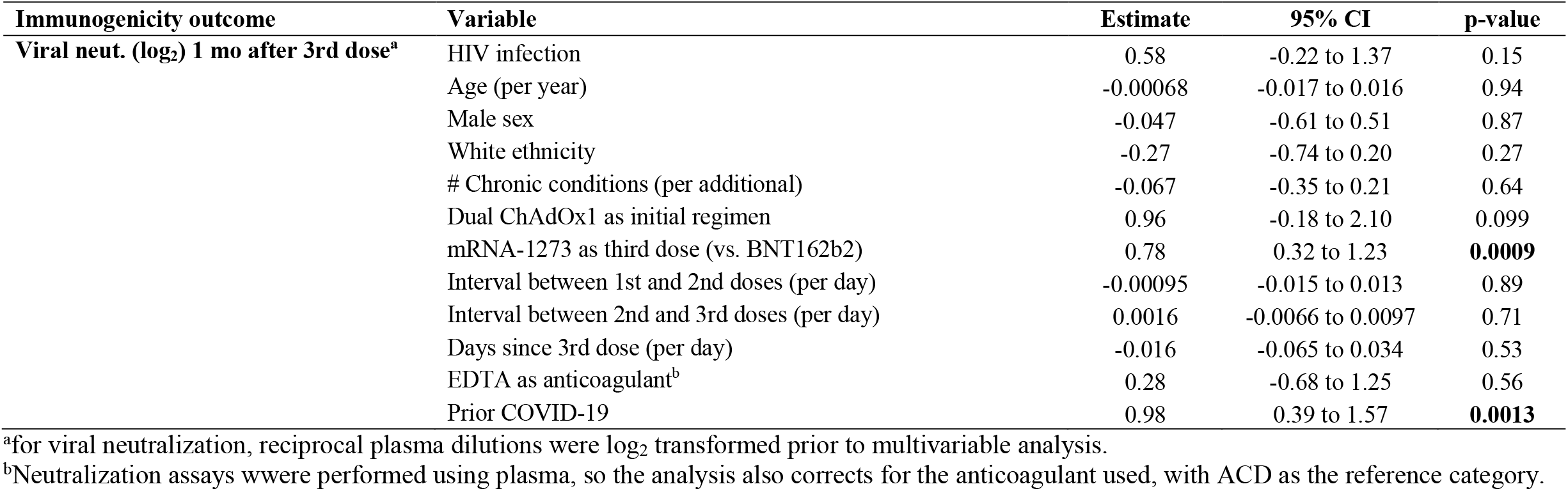
Multivariable analysis of the relationship between sociodemographic, health and vaccine-related variables and viral neutralization activity one month following the third vaccine dose.

| Immunogenicity outcome | Variable | Estimate | 95% CI | p-value |
| --- | --- | --- | --- | --- |
| Viral neut. (log<sub>2</sub>) 1 mo after 3rd dose<sup>a</sup> | HIV infection | 0.58 | -0.22 to 1.37 | 0.15 |
|  | Age (per year) | -0.00068 | -0.017 to 0.016 | 0.94 |
|  | Male sex | -0.047 | -0.61 to 0.51 | 0.87 |
|  | White ethnicity | -0.27 | -0.74 to 0.20 | 0.27 |
|  | # Chronic conditions (per additional) | -0.067 | -0.35 to 0.21 | 0.64 |
|  | Dual ChAdOx1 as initial regimen | 0.96 | -0.18 to 2.10 | 0.099 |
|  | mRNA-1273 as third dose (vs. BNT162b2) | 0.78 | 0.32 to 1.23 | **0.0009** |
|  | Interval between 1st and 2nd doses (per day) | -0.00095 | -0.015 to 0.013 | 0.89 |
|  | Interval between 2nd and 3rd doses (per day) | 0.0016 | -0.0066 to 0.0097 | 0.71 |
|  | Days since 3rd dose (per day) | -0.016 | -0.065 to 0.034 | 0.53 |
|  | EDTA as anticoagulant<sup>b</sup> | 0.28 | -0.68 to 1.25 | 0.56 |
|  | Prior COVID-19 | 0.98 | 0.39 to 1.57 | **0.0013** |

<sup>a</sup>for viral neutralization, reciprocal plasma dilutions were log<sub>2</sub> transformed prior to multivariable analysis.
<sup>b</sup>Neutralization assays were performed using plasma, so the analysis also corrects for the anticoagulant used, with ACD as the reference category.

## REFERENCES

1. Geretti AM, Stockdale AJ, Kelly SH, et al. Outcomes of COVID-19 related hospitalization among people with HIV in the ISARIC WHO Clinical Characterization Protocol (UK): a prospective observational study. Clin Infect Dis 2020.

2. Boulle A, Davies MA, Hussey H, et al. Risk factors for COVID-19 death in a population cohort study from the Western Cape Province, South Africa. Clin Infect Dis 2020.

3. Tesoriero JM, Swain CE, Pierce JL, et al. COVID-19 Outcomes Among Persons Living With or Without Diagnosed HIV Infection in New York State. JAMA network open 2021; 4:e2037069.

4. Bhaskaran K, Rentsch CT, MacKenna B, et al. HIV infection and COVID-19 death: a population-based cohort analysis of UK primary care data and linked national death registrations within the OpenSAFELY platform. Lancet HIV 2021; 8:e24–e32.

5. Poland GA, Ovsyannikova IG, Kennedy RB. SARS-CoV-2 immunity: review and applications to phase 3 vaccine candidates. Lancet 2020; 396:1595–606.

6. Baden LR, El Sahly HM, Essink B, et al. Efficacy and Safety of the mRNA-1273 SARS-CoV-2 Vaccine. N Engl J Med 2021; 384:403–16.

7. Polack FP, Thomas SJ, Kitchin N, et al. Safety and Efficacy of the BNT162b2 mRNA Covid-19 Vaccine. N Engl J Med 2020; 383:2603–15.

8. Massarweh A, Eliakim-Raz N, Stemmer A, et al. Evaluation of Seropositivity Following BNT162b2 Messenger RNA Vaccination for SARS-CoV-2 in Patients Undergoing Treatment for Cancer. JAMA oncology 2021; 7:1133–40.

9. Apostolidis SA, Kakara M, Painter MM, et al. Cellular and humoral immune responses following SARS-CoV-2 mRNA vaccination in patients with multiple sclerosis on anti-CD20 therapy. Nat Med 2021.

10. Moor MB, Suter-Riniker F, Horn MP, et al. Humoral and cellular responses to mRNA vaccines against SARS-CoV-2 in patients with a history of CD20 B-cell-depleting therapy (RituxiVac): an investigator-initiated, single-centre, open-label study. The Lancet Rheumatology 2021.

11. Deepak P, Kim W, Paley MA, et al. Effect of Immunosuppression on the Immunogenicity of mRNA Vaccines to SARS-CoV-2 : A Prospective Cohort Study. Ann Intern Med 2021.

12. Grupper A, Rabinowich L, Schwartz D, et al. Reduced humoral response to mRNA SARS-CoV-2 BNT162b2 vaccine in kidney transplant recipients without prior exposure to the virus. Am J Transplant 2021; 21:2719–26.

13. Plana M, García F, Gallart T, et al. Immunological benefits of antiretroviral therapy in very early stages of asymptomatic chronic HIV-1 infection. Aids 2000; 14:1921–33.

14. Kaufmann GR, Zaunders JJ, Cunningham P, et al. Rapid restoration of CD4 T cell subsets in subjects receiving antiretroviral therapy during primary HIV-1 infection. Aids 2000; 14:2643–51.

15. Bart PA, Rizzardi GP, Tambussi G, et al. Immunological and virological responses in HIV-1-infected adults at early stage of established infection treated with highly active antiretroviral therapy. Aids 2000; 14:1887–97.

16. Lundgren JD, Babiker AG, Gordin F, et al. Initiation of Antiretroviral Therapy in Early Asymptomatic HIV Infection. N Engl J Med 2015; 373:795–807.

17. El Chaer F, El Sahly HM. Vaccination in the Adult Patient Infected with HIV: A Review of Vaccine Efficacy and Immunogenicity. Am J Med 2019; 132:437–46.

18. Kernéis S, Launay O, Turbelin C, Batteux F, Hanslik T, Boëlle PY. Long-term immune responses to vaccination in HIV-infected patients: a systematic review and meta-analysis. Clin Infect Dis 2014; 58:1130–9.

19. Geretti AM, Doyle T. Immunization for HIV-positive individuals. Curr Opin Infect Dis 2010; 23:32–8.

20. Frater J, Ewer KJ, Ogbe A, et al. Safety and immunogenicity of the ChAdOx1 nCoV-19 (AZD1222) vaccine against SARS-CoV-2 in HIV infection: a single-arm substudy of a phase 2/3 clinical trial. Lancet HIV 2021; 8:e474–e85.

21. Madhi SA, Koen AL, Izu A, et al. Safety and immunogenicity of the ChAdOx1 nCoV-19 (AZD1222) vaccine against SARS-CoV-2 in people living with and without HIV in South Africa: an interim analysis of a randomised, double-blind, placebo-controlled, phase 1B/2A trial. Lancet HIV 2021; 8:e568–e80.

22. Levy I, Wieder-Finesod A, Litchevsky V, et al. Immunogenicity and safety of the BNT162b2 mRNA COVID-19 vaccine in people living with HIV-1. Clin Microbiol Infect 2021.

23. Woldemeskel BA, Karaba AH, Garliss CC, et al. The BNT162b2 mRNA Vaccine Elicits Robust Humoral and Cellular Immune Responses in People Living with HIV. Clin Infect Dis 2021.

24. Ruddy JA, Boyarsky BJ, Bailey JR, et al. Safety and antibody response to two-dose SARS-CoV-2 messenger RNA vaccination in persons with HIV. Aids 2021.

25. Noe S, Ochana N, Wiese C, et al. Humoral response to SARS-CoV-2 vaccines in people living with HIV. Infection 2021:1–7.

26. Balcells ME, Le Corre N, Durán J, et al. Reduced immune response to inactivated SARS-CoV-2 vaccine in a cohort of immunocompromised patients in Chile. Clin Infect Dis 2022.

27. Haidar G, Agha M, Bilderback A, et al. Prospective evaluation of COVID-19 vaccine responses across a broad spectrum of immunocompromising conditions: the COVICS study. Clin Infect Dis 2022.

28. Brumme ZL, Mwimanzi F, Lapointe HR, et al. Humoral immune responses to COVID-19 vaccination in people living with HIV receiving suppressive antiretroviral therapy. NPJ vaccines 2022; 7:28.

29. Mizrahi B, Lotan R, Kalkstein N, et al. Correlation of SARS-CoV-2-breakthrough infections to time-from-vaccine. Nat Commun 2021; 12:6379.

30. Goldberg Y, Mandel M, Bar-On YM, et al. Waning Immunity after the BNT162b2 Vaccine in Israel. N Engl J Med 2021; 385:e85.

31. Levin EG, Lustig Y, Cohen C, et al. Waning Immune Humoral Response to BNT162b2 Covid-19 Vaccine over 6 Months. N Engl J Med 2021; 385:e84.

32. Doria-Rose NA, Shen X, Schmidt SD, et al. Booster of mRNA-1273 Vaccine Reduces SARS-CoV-2 Omicron Escape from Neutralizing Antibodies. medRxiv 2021.

33. Garcia-Beltran WF, St Denis KJ, Hoelzemer A, et al. mRNA-based COVID-19 vaccine boosters induce neutralizing immunity against SARS-CoV-2 Omicron variant. medRxiv 2021.

34. Schmidt F, Muecksch F, Weisblum Y, et al. Plasma neutralization properties of the SARS-CoV-2 Omicron variant. medRxiv 2021.

35. Collie S, Champion J, Moultrie H, Bekker LG, Gray G. Effectiveness of BNT162b2 Vaccine against Omicron Variant in South Africa. N Engl J Med 2021.

36. Cele S, Jackson L, Khan K, et al. SARS-CoV-2 Omicron has extensive but incomplete escape of Pfizer BNT162b2 elicited neutralization and requires ACE2 for infection. medRxiv 2021.

37. Ogbe A, Pace M, Bittaye M, et al. Durability of ChAdOx1 nCov-19 (AZD1222) vaccination in people living with HIV - responses to SARS-CoV-2, variants of concern and circulating coronaviruses. medRxiv 2021:2021.09.28.21264207.

38. Bar-On YM, Goldberg Y, Mandel M, et al. Protection of BNT162b2 Vaccine Booster against Covid-19 in Israel. N Engl J Med 2021; 385:1393–400.

39. Choi A, Koch M, Wu K, et al. Safety and immunogenicity of SARS-CoV-2 variant mRNA vaccine boosters in healthy adults: an interim analysis. Nat Med 2021; 27:2025–31.

40. Falsey AR, Frenck RW, Jr., Walsh EE, et al. SARS-CoV-2 Neutralization with BNT162b2 Vaccine Dose 3. N Engl J Med 2021; 385:1627–9.

41. Mattiuzzo G, Bentley EM, Hasall M, et al. Establishment of the WHO International Standard and Reference Panel for anti-SARS-CoV-2 antibody. In: WHO_Expert_Committee_on_Biological_Standardization, ed. Vol. WHO/BS/2020.2402. Geneva: World_Health_Organization, 2020.

42. Tan CW, Chia WN, Qin X, et al. A SARS-CoV-2 surrogate virus neutralization test based on antibody-mediated blockage of ACE2-spike protein-protein interaction. Nat Biotechnol 2020; 38:1073–8.

43. Notarte KI, Guerrero-Arguero I, Velasco JV, et al. Characterization of the significant decline in humoral immune response six months post-SARS-CoV-2 mRNA vaccination: A systematic review. J Med Virol 2022.

44. Collie S, Champion J, Moultrie H, Bekker LG, Gray G. Effectiveness of BNT162b2 Vaccine against Omicron Variant in South Africa. N Engl J Med 2022; 386:494–6.

45. Cele S, Jackson L, Khoury DS, et al. Omicron extensively but incompletely escapes Pfizer BNT162b2 neutralization. Nature 2022; 602:654–6.

46. Garcia-Beltran WF, St Denis KJ, Hoelzemer A, et al. mRNA-based COVID-19 vaccine boosters induce neutralizing immunity against SARS-CoV-2 Omicron variant. Cell 2022; 185:457–66.e4.

47. Planas D, Saunders N, Maes P, et al. Considerable escape of SARS-CoV-2 Omicron to antibody neutralization. Nature 2022; 602:671–5.

48. Keeton R, Tincho MB, Ngomti A, et al. T cell responses to SARS-CoV-2 spike cross-recognize Omicron. Nature 2022.

49. Liu J, Chandrashekar A, Sellers D, et al. Vaccines elicit highly conserved cellular immunity to SARS-CoV-2 Omicron. Nature 2022.

50. Feng S, Phillips DJ, White T, et al. Correlates of protection against symptomatic and asymptomatic SARS-CoV-2 infection. Nat Med 2021; 27:2032–40.

